# A comprehensive framework for the interpretation of *TTN* missense variants

**DOI:** 10.1101/2025.07.17.25331109

**Authors:** Maria Francesca Di Feo, Martin Rees, Victoria Lillback, Ay Lin Kho, Angelina Meybatova, Mark Holt, Heinz Jungbluth, Francesco Muntoni, Giovanni Baranello, Anna Sarkozy, Chiara Fiorillo, Serena Baratto, Claudio Bruno, Monica Traverso, Michele Iacomino, Marina Pedemonte, Noemi Brolatti, Francesca Faravelli, Federico Zara, G.M. Luana Mandarà, Alan H. Beggs, Casie A. Genetti, Pamela Barraza-Flores, Carmelo Rodolico, Sonia Messina, Franziska Schnabel, Istvan Balogh, Katalin Szakszon, Siiri Sarv, Katrin Õunap, Federica Silvia Ricci, Alessandro Mussa, Edoardo Malfatti, Enrico Silvio Bertini, Adele D’Amico, Daria Diodato, Michela Catteruccia, Gianina Ravenscroft, Mridul Johari, Sergei A. Kurbatov, Polina Chausova, Aysylu Murtazina, Anna Kuchina, Olga Shchagina, Minas Drakos, Martha Spilioti, Athanasios E. Evangeliou, Ioannis Zaganas, Huahua Zhong, Sushan Luo, Luciano Merlini, Cam-Tu-Emilie Nguyen, Giorgio Tasca, Tara Reeves, Stellan Mörner, Olof Danielsson, Bjarne Udd, TTN study group, Mathias Gautel, Marco Savarese

**Affiliations:** Folkhälsan Research Center, Helsinki, Finland; Department of Medical Genetics, Medicum, University of Helsinki, Haartmaninkatu 8, Helsinki, 00290, Finland; Department of Neuroscience, Rehabilitation, Ophthalmology, Genetics, and Maternal and Child Health (DINOGMI), University of Genoa, Genova, Italy; Randall Centre for Cell & Molecular Biophysics, School of Basic & Medical Biosciences, King’s College London, London SE1 1UL, United Kingdom; Children’s Neuroscience Centre, Evelina London Children’s Hospital, Guy’s and St. Thomas’ Hospital NHS Foundation Trust, London, United Kingdom; The Dubowitz Neuromuscular Unit, UCL Great Ormond Street Institute of Child Health, NIHR Great Ormond Street Hospital Biomedical Research Centre and Great Ormond Street Hospital NHS Foundation Trust, London, UK; Child Neuropsychiatry Unit, IRCCS Istituto Giannina Gaslini, Genoa, Italy; Centre of Translational and Experimental Myology, IRCCS Istituto Giannina Gaslini, Genoa, Italy; Medical Genetics Unit, IRCCS Giannina Gaslini Hospital, Genova, Italy; Pediatric Neurology and Muscle Diseases Unit, IRCCS Giannina Gaslini Hospital, Genova, Italy; Clinical Genetics and Genomics Unit, IRCCS Giannina Gaslini Hospital, Genova, Italy; Maria Paternò Arezzo Hospital, Ragusa, Italy; Division of Genetics and Genomics, The Manton Center for Orphan Disease Research, Boston Children’s Hospital, Harvard Medical School, Boston, MA, USA; Department of Clinical and Experimental Medicine, University of Messina, Italy; University of Leipzig Medical Center, Leipzig, Germany; University of Debrecen, Faculty of Medicine, Department of Medical Genetics, Debrecen, Hungary; University of Debrecen, Faculty of Medicine, Institute of Paediatrics, Debrecen, Hungary; Genetic and Personalized Medicine Clinic, Tartu University Hospital and Institute of Clinical Medicine, University of Tartu, Tartu, Estonia; Department of Public Health and Pediatrics, University of Turin, Regina Margherita Children’s Hospital, Torino, Italy; Université Paris Est Créteil, Inserm U955, IMRB, Reference Center for Neuromuscular Disorders, APHP Henri Mondor University Hospital, Créteil, France; Research Unit of Neuromuscular and Neurodegenerative Disease, Bambino Gesù Children’s Hospital, IRCCS, Roma, Italy; Harry Perkins Institute of Medical Research, Nedlands, Australia; Voronezh State Medical University, Voronezh, Russian Federation; Research Centre for Medical Genetics, Moscow, Russia; Neurology/Neurogenetics Laboratory, Medical School, University of Crete; 1st Neurology Department, AHEPA University Hospital, Thessaloniki, Greece; Division of Child Neurology, St. Luke’s Hospital, Thessaloniki, Greece; Department of Neurology, University Hospital of Heraklion, Heraklion, Greece; Huashan Rare Disease Centre and Department of Neurology, Huashan Hospital, Shanghai Medical College, National Centre for Neurological Disorders, Fudan University, Shanghai, 200040, China; Department of Biomedical and Neuromotor Science, DIBINEM, University of Bologna, 40136 Bologna, Italy; CHU Sainte-Justine, Montreal, QC, Canada; The John Walton Muscular Dystrophy Research Centre, Translational and Clinical Research Institute, Newcastle University and Newcastle Hospitals NHS Foundation Trust, Newcastle upon Tyne NE1 3BZ, UK; Department of Public Health and Clinical Medicine, Umeå University, Umeå, Sweden; Division of Neurology, Department of Biomedical and Clinical Sciences, Faculty of Medicine and Health Sciences, Linköping University, Linköping, Sweden; Neuromuscular Research Center, Tampere University and University Hospital, Tampere, Finland; Neuropaediatrics Department, Hospital Sant Joan De Déu, Institut De Recerca Sant Joan De Déu, Barcelona, 08950 Spain; Neuromuscular Unit, Department of Neurology, Hospital Sant Joan De Déu, Barcelona, Spain; Neuropathology Lab, Neurosciences Centre, All India Institute of Medical Sciences, New Delhi, India; Department of Neurology, All India Institute of Medical Sciences, New Delhi, India; CSIR – Institute of Genomics and Integrative Biology, New Delhi, India; Department Pediatric Neurology, All India Institute of Medical Sciences, New Delhi, India; Centre of Molecular Biology and Genetics, Brno University Hospital, Brno, Czech Republic

**Keywords:** Titin, titinopathies, missense variants, functional evidence, in silico tools, AlphaMissense

## Abstract

**Background:** Missense variants in *TTN* pose a major challenge in genetic diagnostics due to their high frequency in the general population, the large size of the gene, and the complex multidomain architecture of the titin protein. While the contribution of truncating variants (TTNtv) to titinopathies is well established, the role of rare *TTN* missense variants remains poorly defined. Advances in computational prediction and functional testing offer new tools to assess their potential pathogenicity, which however are currently not fully utilized for clinical application.

**Methods:** We analyzed an international cohort of unsolved myopathy cases selected based on the presence of a rare missense variant in trans with a TTNtv. Clinical data were collected from neuromuscular centers worldwide. In silico predictions were generated using AlphaMissense and complemented by MAF and exon usage information. Additional inclusion criteria were based on a Minor Allele Frequency < 0.010 and an AlphaMissense score ≥ 0.792 for the missense variant, in accordance with the latest ClinGen guidelines. Selected missense variants were characterized in vitro through protein expression and cell imaging assays to assess their effects on domain solubility and aggregation.

**Results:** Thirty patients with TTNtv/missense combinations were identified, presenting with heterogeneous myopathic phenotypes, ranging from congenital to adult onset. An in-depth analysis on AlphaMissense predictions highlighted those changes most frequently predicted as possibly pathogenic. Functional assays showed that three selected variants with changes to proline, located in β-sheets of Ig domains, led to impaired folding, cytoplasmic aggregation and co-localisation with proteostasis markers. In our cohort, all non-proline mutations occurred at buried sites, while some proline substitutions affected exposed residues. Notably, the variant p.(Gln7023Pro) was identified in 5 unrelated families sharing a conserved haplotype, indicating a common ancestor. This variant and the previously reported p.(Arg25480Pro) variant have been reclassified as likely pathogenic.

**Conclusions:** By integrating clinical, computational, and functional evidence, we propose a framework for interpreting *TTN* missense variants. Combining multiple lines of evidence is essential for variants’ classification and interpretation, especially given *TTN* complexity. Advancing diagnostic accuracy will require tailored interpretation guidelines and a global effort in data sharing and functional validation.

## Background

*TTN* is a 294-kb gene of 364 exons that encodes the protein titin, the largest protein in the human body and among the largest proteins known in nature.^1^ Titin is expressed in striated muscle and spans half a sarcomere, the basic contractile unit of muscle, from the Z-disk to the M-band (Figure 1). The protein plays roles in muscle development, structure, and stabilization and is composed of 132 fibronectin III-like (Fn3) domains, up to 169 immunoglobulin-like (Ig) domains, a single kinase domain, and unstructured regions.^2,3^ Each region of titin has specific features and functions: the Z-disk-anchored region links titin to the actin-based thin filament via alpha-actinin; the I-band, including the segmental duplication region of *TTN* spanning from exon 173 to exon 199, plays a crucial role in muscle stiffness and elasticity; the A-band region helps assemble and regulate the length of the myosin-based thick filaments whilst, finally, the M-band region contributes to sarcomere structural integrity and myofibril formation while interacting with several ligands.^4^ The length and composition of *TTN* regions vary between isoforms, which are differentially expressed depending on the stage of development and type of muscle.^5,6^ The A-band is the most constitutively expressed region, with minimal exon usage variation reported across major transcripts. In contrast, other regions contain exons with variable percentage spliced-in (PSI) values.^6,7^

**Figure 1.**
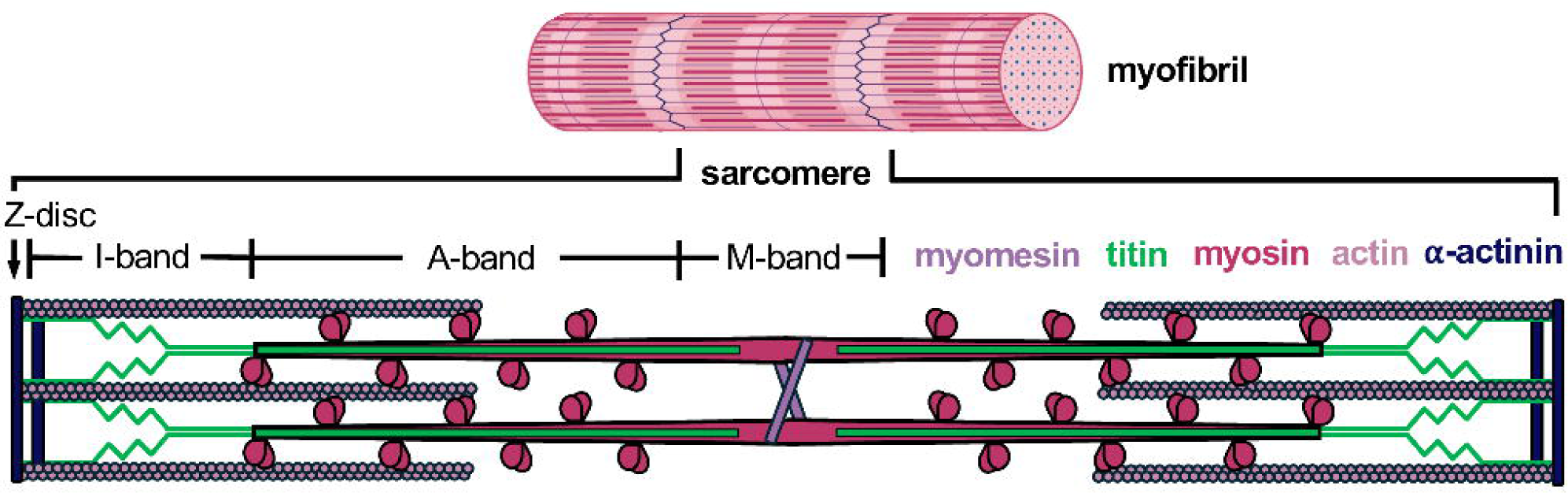
Schematic representation of the sarcomere, illustrating its main structural regions (Z-disk, I-band, A-band, M-band) and key structural proteins.

Pathogenic *TTN* variants cause a wide range of skeletal myopathies, cardiomyopathies or combinations of both, varying by their mode of inheritance (autosomal dominant, autosomal recessive, compound heterozygous or digenic), age of onset, muscle involvement, severity, and rate of progression.^8–12^ Pathogenic missense variants in two specific exons, 344 and 364, cause hereditary myopathy with early respiratory failure (HMERF) and tibial muscular dystrophy (TMD or Udd myopathy), respectively, with the variants shown to render their domain insoluble or unfolded when expressed in bacteria, or to reduce the thermal stability of the domain.^10,13,14^ Heterozygous *TTN* truncating variants (*TTN*tv) in exons with high PSI values – representing the proportion of transcripts in which the corresponding exon is included – in heart muscles is observed in approximately 20% of all dilated cardiomyopathy (DCM) cases and are thought to contribute to disease by causing haploinsufficiency and/or resulting in a poison peptide.^15–17^

Most of *TTN*-related skeletal myopathies identified to date are linked to biallelic *TTN*tv.^18^ Population studies have found that up to 1% of the general population carries a *TTN*tv, and up to 20% carry a rare missense variant with a Minor Allele Frequency (MAF) <1%.^19,20^ Recently, there has been increased focus on the investigation of rare missense variants throughout *TTN*; with the exception of pathogenic dominant variants in exons 344 and 364, however, only a few studies have addressed this issue, and fewer than a dozen missense variants linked to disease reported — mainly in sporadic cases — and with limited supporting evidence for their pathogenicity according to the criteria established by the American College of Medical Genetics and Genomics (ACMG).^21–23^ Clinical interpretation of *TTN* missense variants is complex, given the large size of the gene, the frequency of the variants, and the default application of the BP1 benign “supporting” criterion (“missense variant in a gene for which primarily truncating variants are known to cause disease”).^23^ Our study represents an international effort to collect unsolved cases with a suspected recessive titinopathy to gather further evidence on rare *TTN* missense variants most likely affecting the protein function or structure. We hypothesize that missense changes in *TTN* may represent a major but currently frequently overlooked contributor to inherited titinopathies.

## Methods

### In silico prediction and variants assessment

We collected all the available predictions for missense variants in the *TTN* gene generated by AlphaMissense, the state-of-the-art artificial intelligence model developed to assess the pathogenic potential of missense variants by integrating sequence context, evolutionary conservation, and other biological features.^24^ As a cut-off for AlphaMissense predictions—which range from 0 to 1, with higher scores indicating a greater likelihood of pathogenicity—we applied the thresholds proposed in the updated ClinGen recommendations for PP3/BP4 variant classification criteria.^25^ In this recent work, Bergquist and colleagues assessed the validity of the most recent in silico tools to support a deleterious effect on the gene or gene product. After posterior probability-based calibration, AlphaMissense reached the Strong level of evidence for pathogenicity (PP3) and at least the Moderate level for benignity (BP4), although at score thresholds more stringent than those originally recommended by its developers. We therefore applied the following recommended thresholds: ≤ 0.070 for “Benign” BP3 (–3 points); [0.071, 0.099] for “Benign Moderate” BP3 (–2 points); [0.100, 0.169] for “Benign Supporting” (− 1 point); [0.170, 0.791] for “Indeterminate”; [0.792 – 0.905] for “Pathogenic Supporting” PP3 (+ 1 point); [0.906 – 0.971] for “Pathogenic Moderate” PP3 (+2 points); [0.972, 0.989] for “Pathogenic” PP3 (+ 3 points); ≥ 0.990 for “Pathogenic Strong” (+ 4 points). Together with AlphaMissense predictions, we reported available data on Minor Allele Frequency (MAF) for each *TTN* missense variants from the gnomAD database (Additional File 1).^26^ For each variant, whether TTNtv or missense, the PSI of the corresponding exon was assessed. PSI values—derived from our recent study (open-access database: http://psivis.it.helsinki.fi:3838/TTN_PSIVIS/)—are reported in Table 1 and Table 2.^7^

**Table 1.** Clinical details of patients carrying missense variants to proline (MAF < 0.010, AlphaMissense score ≥ 0.792) that have been functionally assessed in this study, in trans with a TTNtv.

| Onset | Family ID (Patient ID) | Missense variant, position and skeletal muscle PSI | Truncating variant, position and skeletal muscle PSI | Short clinical report |
| --- | --- | --- | --- | --- |
| Congenital onset | F1<br>(F1-P1<br>F1-P2) | **c.21068A>C**<br>**p.(Gln7023Pro)**<br>**exon 73,**<br>**proximal I-band**<br>prenatal/postnatal skeletal muscle PSI: 96%/88% | c.24729C>A<br>p.(Cys8243Ter)<br>exon 86<br>proximal I-band<br>prenatal/postnatal skeletal muscle PSI: 94%/86% | Congenital myopathy, pectus excavatum, scoliosis, scapular winging, respiratory insufficiency, myopathic facies, underweight |
|  | F2<br>(F2-P1) |  | c.55351C>T<br>p.(Arg18451Ter)<br>exon 287<br>A-band<br>prenatal/postnatal skeletal muscle PSI: 94%/87% | Congenital myopathy, clubfeet detected prenatally, contractures at birth, hypotonia, scoliosis, scapular winging, respiratory insufficiency, myopathic facies |
|  | F3<br>(F3-P1) |  | c.68575_68576dup<br>p.(Ile22861SerfsTer14)<br>exon 324<br>A-band<br>prenatal/postnatal skeletal muscle PSI: 93%/86% | Congenital myopathy, scoliosis, respiratory insufficiency |
|  | F4<br>(F4-P1) |  | c.61876C>T<br>p.(Arg20626Ter)<br>exon 305<br>A-band<br>prenatal/postnatal skeletal muscle PSI: 96%/91% | Congenital myopathy, scoliosis, respiratory insufficiency, myopathic facies, underweight, waddling gait, difficulty climbing stairs |
|  | F5<br>(F5-P1) |  | c.26173_26174delAC<br>p.(Thr8725LeufsTer19)<br>exon 91<br>proximal I-band<br>prenatal/postnatal skeletal muscle PSI: 95%/85% | Congenital myopathy, hypotonia, difficulty climbing stairs, difficulty running |
|  | F6<br>(F6-P1) |  | c.32092C>T<br>p.(Arg10698Ter)<br>exon 126 | Congenital myopathy, classified as multiminicore myopathy, hypotonia, scoliosis s/p surgical repair, difficulty climbing stairs, |
|  | (BOS169<br>3-1)<sup>a</sup> |  | I-band<br>prenatal/postnatal<br>skeletal muscle PSI:<br>89%/70% | cannot run. |
| Adult onset | F7<br>(F7-P1) | **c.14486A>C**<br>**p.(Gln4829Pro)**<br>**exon 51,**<br>**proximal I-band**<br>prenatal/postnatal<br>skeletal muscle<br>PSI: 97%/94% | c.42472dup<br>p.(Thr14158AsnfsTer7)<br>exon 232<br>A-band<br>prenatal/postnatal<br>skeletal muscle PSI:<br>97%/91% | Noncongenital distal myopathy |
|  | F8<br>(F8-P1<br>F8-P2) |  | c.26028del<br>p.(Trp8676CysfsTer16)<br>exon 91<br>I-band<br>prenatal/postnatal<br>skeletal muscle PSI:<br>95%/85% | Noncongenital myopathy, proximal with facial<br>involvement |
| Congenital onset | F9<br>(F9-P1) | **c.25039T>C**<br>**p.(Ser8347Pro)**<br>**exon 87,**<br>**proximal I-band**<br>prenatal/postnatal<br>skeletal muscle<br>PSI:<br>94%/86% | c.76109_76110delTA,<br>p.(Ile25370ArgfsTer6)<br>exon 327<br>A-band<br>prenatal/postnatal<br>skeletal muscle PSI:<br>97%/93% | Congenital myopathy, contractures at birth,<br>“frog-like” position, dysphagia in infancy<br>(resolved), Bi-PAP home ventilation support,<br>scoliosis, pectus excavatum, mild facial<br>dysmorphisms (mandibular prognathism,<br>maxillary hypoplasia) |

**Table 2.**
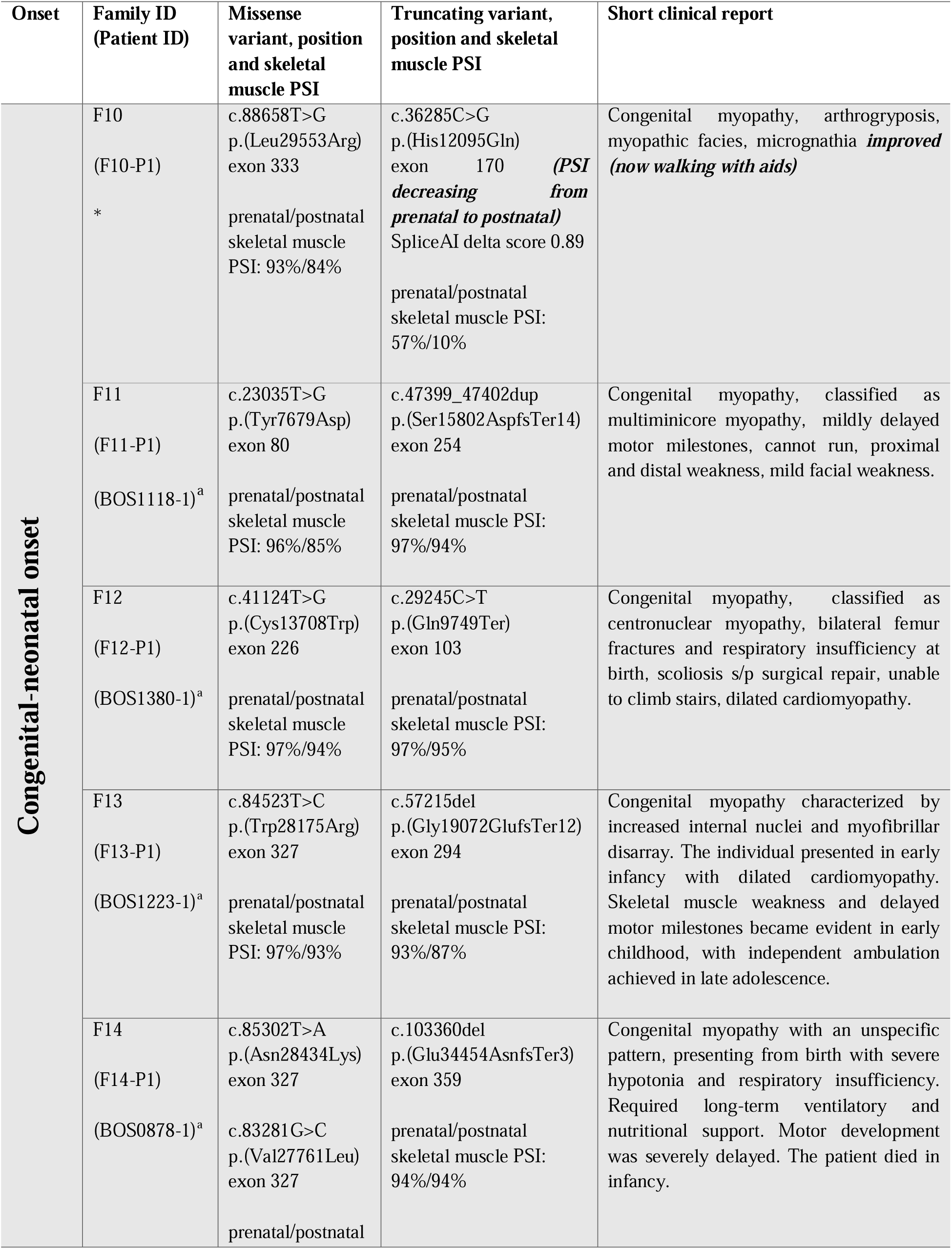
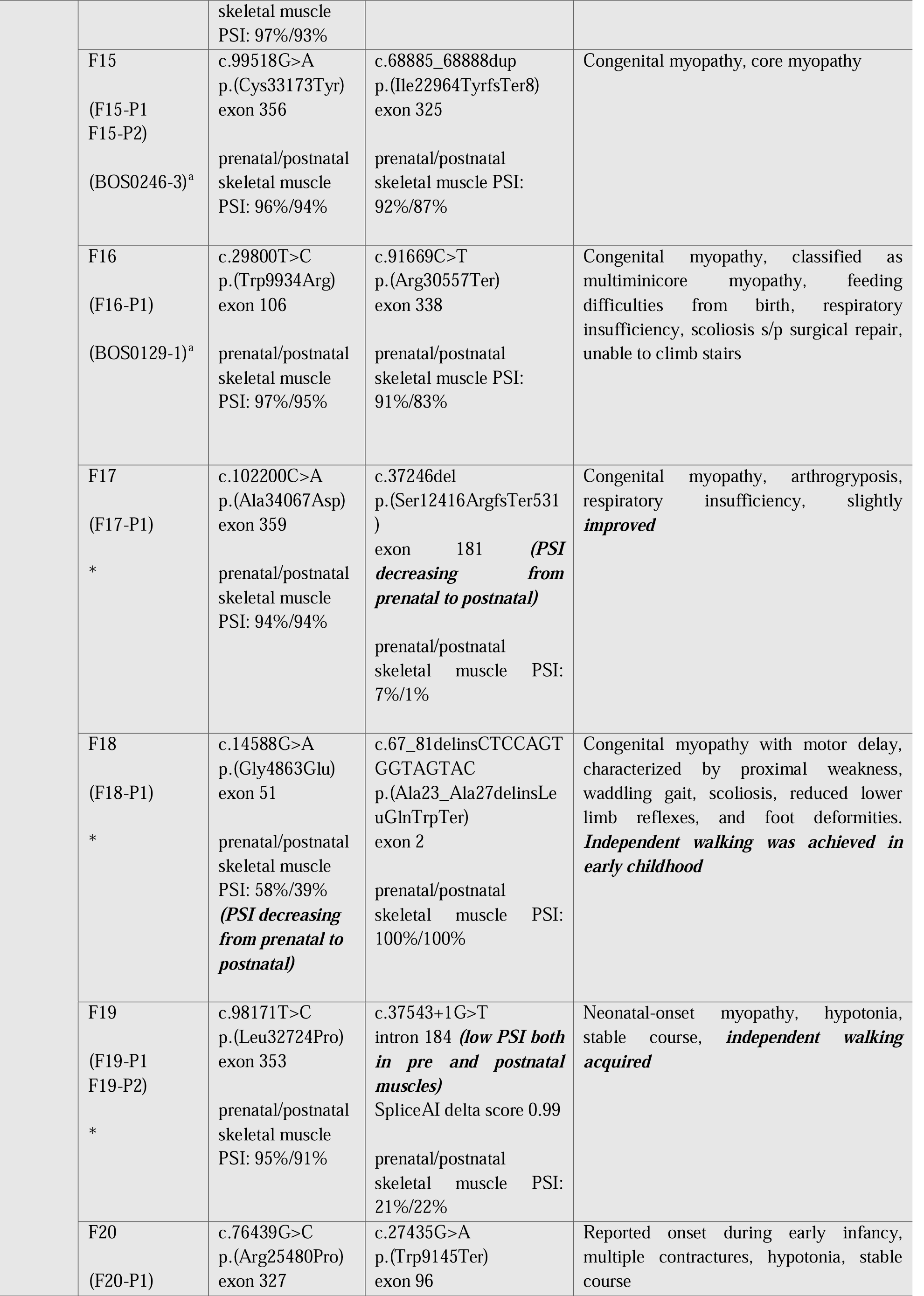
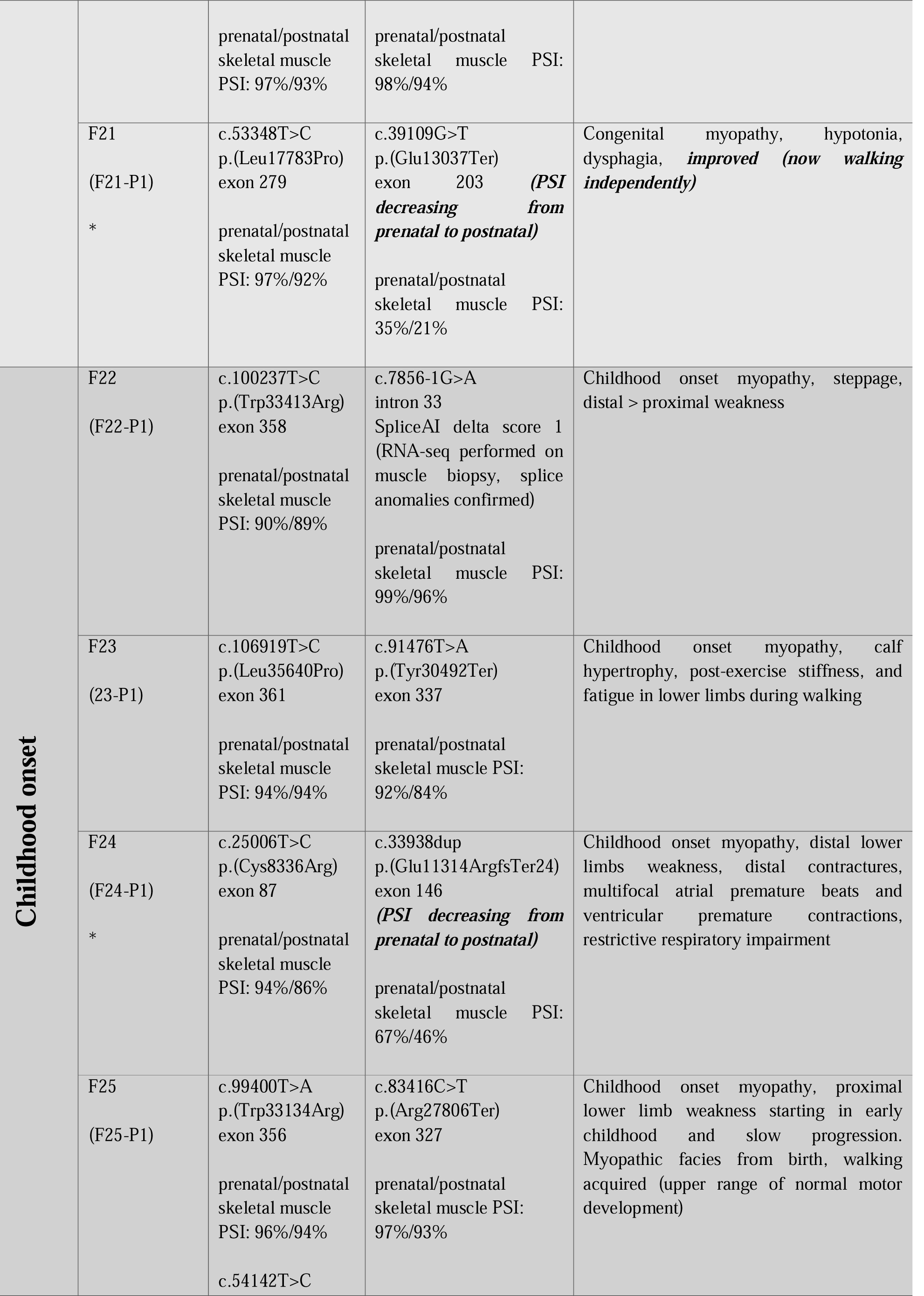
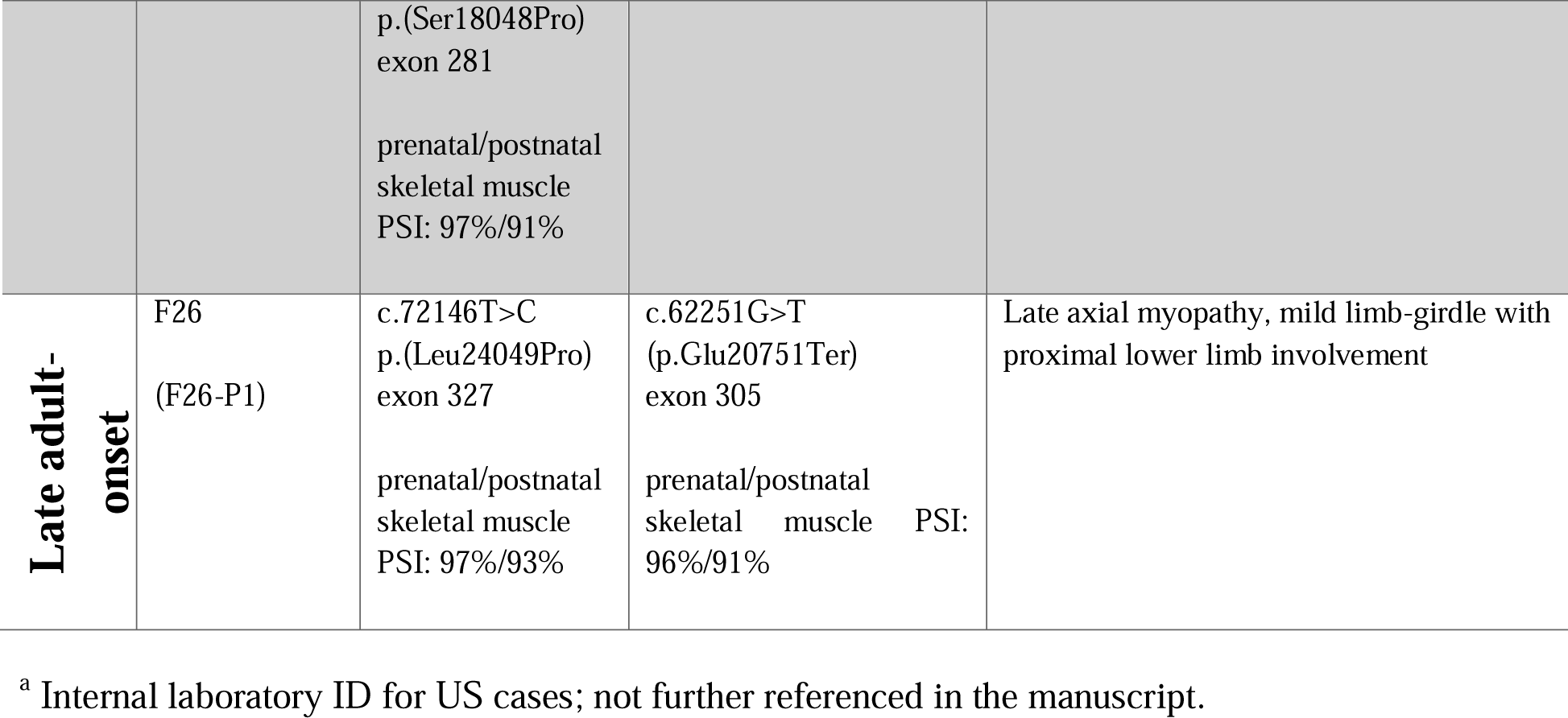
Clinical details of patients carrying missense variants (MAF < 0.010, AlphaMissense score ≥ 0.792), not functionally assessed in this study, in trans with a TTNtv.

### Data collection

We collected from several international neuromuscular centers *TTN* data from unsolved myopathy cases carrying a heterozygous likely pathogenic/pathogenic TTNtv, in compound heterozygosity with a rare missense variant. Only cases with missense variants showing a MAF <0.010 and an AlphaMissense score ≥ 0.792 were included in this study.^27^ All these patients had undergone exome/genome sequencing in their reference hospitals or as part of a resequencing project for unsolved cases, and did not carry other candidate variants with a compatible mode of inheritance (Likely Pathogenic/Pathogenic according to ACMG criteria). CNV analysis was also performed using standard computational tools integrated into the respective diagnostic pipelines. In addition, RNA sequencing (RNA-seq) was performed in two cases, and long-read sequencing in another. We excluded from the current study patients with missense variants in exons 344 and 364, as pathogenic missense variants in these exons have already been associated with specific phenotypes.^10,13^ All variants were reassessed using SpliceAI and Varsome; no evidence of splicing effects was found for variants classified as missense. In case of splicing variants found in trans with a missense variant, SpliceAI scores have been specified in Table 1 and Table 2; all reported scores are above 0.8.^28^ For each potential amino acid substitution, we assessed key parameters including: the pathogenicity score from AlphaMissense, the recurrence of the variant across unrelated families, and the availability of functional evidence. Detailed phenotypic description and age at onset and at last examination were also collected. Biopsies, MRI reports, and clinical photographs were collected where possible (Figure S1-S3, Additional File 2; Additional data and materials available upon reasonable request).

### Bioinformatics

Titin domain boundaries were based on those from TITINdb.^29,30^ Alphafold-predicted structures for the Ig-31 and Ig-54 domains were retrieved from TTITINdb, and the Ig-68 domain was from the experiential structure 3B43; all were visualized using PyMol (Schrödinger Inc.).^31^ The surface accessible area of each wild type (WT) residue of interest mutated in patients was calculated using POPSCOMP data retrieved from TITINdb.^29^

### Cloning, protein expression and cell fractionation

Sequences encoding titin domains Ig-31, Ig-54 and Ig-68 (whose structures are shown in Figure S4, Additional File 2) were amplified from a human skeletal muscle cDNA library by polymerase chain reaction and inserted into a modified pET-14 vector encoding an N-terminal His_6_-tag. Once vectors encoding the WT sequences were obtained, mutations encoding the missense variants Q4829P, Q7023P, and S8347P were induced by mutagenesis PCR. From these vectors, the sequences were amplified and inserted into pCMV_GFPC2 vectors. All vector sequences were confirmed by Sanger sequencing. The pET vectors were transformed into BL21-Star (Thermo Fisher) and cultured overnight in LB broth supplemented with carbenicillin (100ug/mL) at 37°C. 80ul overnight culture was used to inoculate 4mL MagicMedia (Thermo Fisher) which was cultured at 37°C for 8 hours and then 18°C for 48 hours. Cultures were harvested by centrifugation and stored at –80°C.

For Western blot solubility analysis, the cell pellet was resuspended in 20mM HEPES pH7.5, 300mM NaCl and 15mM imidazole, supplemented with cOmplete protease inhibitor (Roche) and then lysed with B-Per (Thermo Fisher) supplemented with lysozyme (0.1mg/mL) and Benzonase (40U/mL, Merck) for 30 minutes at room temperature. 20ul of this sample was mixed with 20ul SDS-PAGE loading buffer for the “total” fraction, and then the lysed cells were separated into soluble and insoluble fractions by centrifugation of 1000g RCF at 4°C for 30 minutes. 20ul of the supernatant was then mixed with 20ul SDS-PAGE loading buffer for the “soluble” fraction. The “total” and “soluble” samples were then incubated at 98°C for 10 minutes.

### Western blotting of protein fragments

10ul of each sample were loaded onto a precast 4-20% polyacrylamide gel (BioRad) and electrophoresis was run until the dye front reached the bottom of the gel. Following electrophoresis, proteins were transferred to nitrocellulose membrane (GE), stained with Ponceau S and imaged. The membrane was then blocked in 5% milk in low-salt buffer (LSB) for 1 hour and incubated in primary antibody (anti-His tag, Novagen, 1:2000 dilution) overnight at 4°C. After washing in LSB, the membrane was incubated in secondary antibody (HRP-tagged anti-mouse, DAKO, 1:1000 dilution) for 1 hour and then washed again. HRP activity was measured following incubation with 2ml Clarity ECL substrate (BioRad) using a ChemDoc (BioRad).

### COS-7 transfection, fixation, and immunostaining

COS-7 cells were prepared using standard protocols and plated at a cell density of 7.5 × 10^4 cells per well of a 24-well plate (Azenta). Vectors encoding GFP-tagged WT Ig-31, Ig-54, and Ig-68 and their variants were transfected using Escort IV (Sigma, 1.7ul per well, added to 0.3ug DNA) according to the manufacturer’s instructions. Following incubation for 24 hours at 37°C, 5% CO_2_, cells were fixed by washing in PBS, incubated with 4% PFA for 10 minutes, and finally washed again in PBS. A subset of cells were permeabilised with 10mg/ml digitonin (30mins) before being blocked with 5% normal goat serum in antibody dilution buffer (1% bovine serum albumin, 1mM Tris pH 7.5, 15.5mM NaCl, 0.2mM EGTA, 0.2mM MgCl_2_, 30mins) and incubated with a primary antibody against conjugated ubiquitin (1:100, FK2, 04-263, Merck, overnight at 4°C) followed by Cy3-conjugated anti-mouse IgG secondary antibody (1:100 Jackson Immunoresearch, 1hr at RT), and DAPI.

### Cell imaging

Widefield images for fluorescence intensity quantification: Fixed cells expressing GFP-tagged titin domains were imaged using a 10x objective on a fluorescence widefield microscope (Leica). Confocal images were obtained using a 60x oil immersion objective on a Nikon AXR inverted confocal microscope (Nikon Imaging Centre, King’s College London).

### Image analysis

Cell expression patterns in widefield fluorescence microscope images were quantified by measuring the Jaccard Index of nuclear overlap with the whole cell mask using two different filters to segment the images into binary masks. Nuclei were identified by their size and roundness. Image segmentation and analysis were performed using a custom script written in Wolfram Mathematica 13.3 (Champaign, Il).

### Statistics

Statistical differences between Jaccard Index values from the analysed images for WT-variant domain pairs were calculated using the Student’s t-test.

## Results

### Overall AlphaMissense predictions on TTN

Of the 224,638 possible amino acid changes corresponding to single nucleotide variants and predicted by AlphaMissense, a total of 47,562 variants is predicted as possibly pathogenic (AlphaMissense score ≥ 0.792, corresponding to PP3 evidence with at least ‘upportin’ strength), as outlined in Figure 2A. Of these, 44,476 (93.5%) are absent from gnomAD. In contrast, less than 10% of missense variants with a MAF > 0.0001 have pathogenic AlphaMissense predictions.

**Figure 2.**
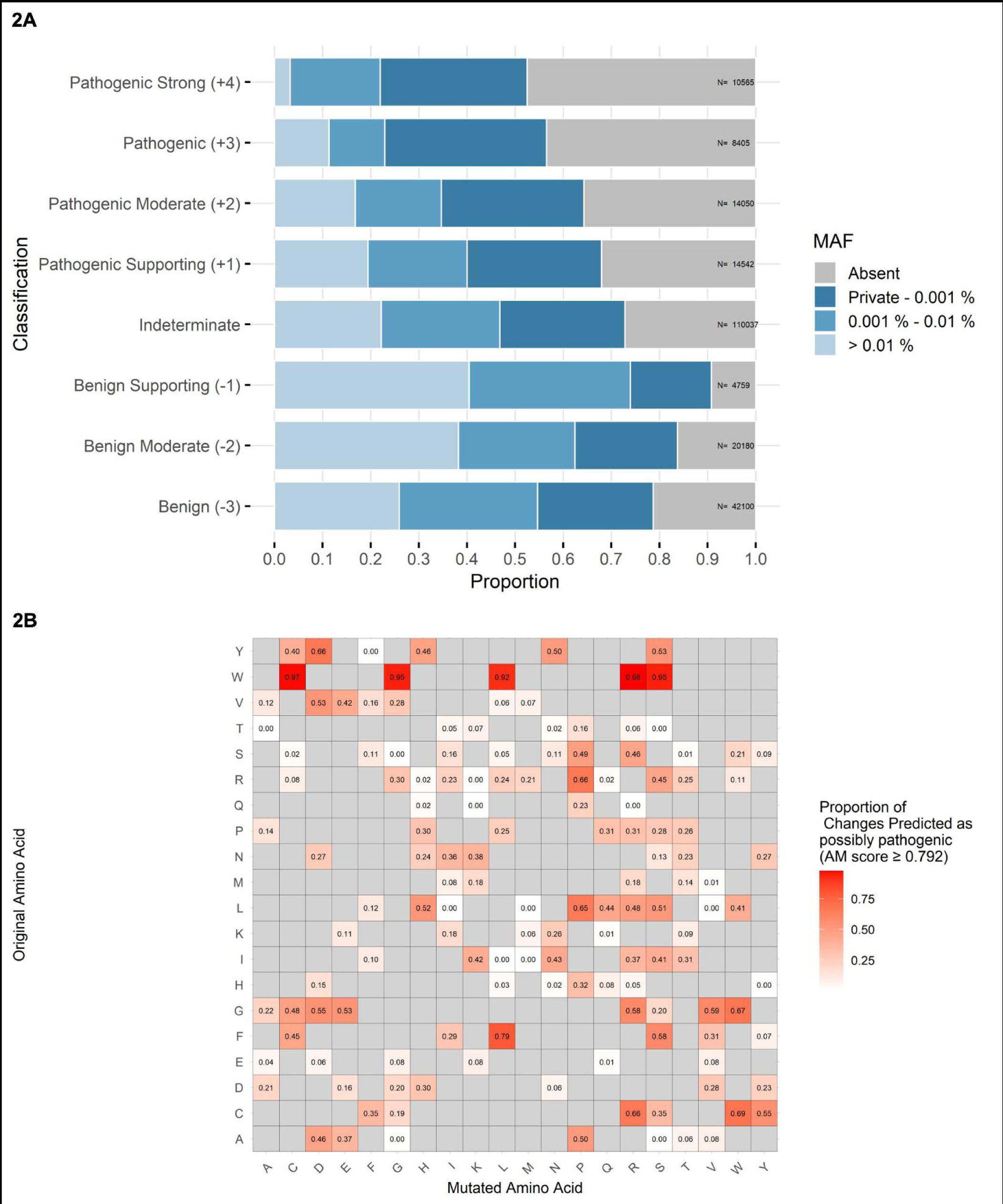
AlphaMissense predictions for *TTN* missense variants. **(A)** Distribution of AlphaMissense prediction scores across minor allele frequency (MAF) categories for TTN missense variants. Variants are grouped by predicted pathogenicity strength, from “Benign” (−3) to “Pathogenic Strong” (+4). **(B)** Heatmap showing the proportion of amino acid substitutions in TTN predicted as possibly pathogenic (Pathogenic Supporting or stronger evidence, AM score ≥ 0.792), with enrichment for specific changes.

Our analysis also identified the amino acid substitutions most frequently predicted as pathogenic by AlphaMissense. The most frequently damaging change is from tryptophan: over 92% of all amino acid changes from tryptophan to another amino acid are predicted to be possibly pathogenic. Titin contains 466 tryptophan residues, from which 3,262 possible missense changes are predicted. Regarding other amino acids, the proportion of damaging changes depends on the specific combinations; e.g. changes to proline, cysteine, arginine, and asparagine are also frequently predicted to be pathogenic, but with greater variability according to the original amino acid (Figure 2B).

### Molecular and clinical findings

We selected 30 unsolved myopathy patients from 26 different families with both a *TTN*tv and a rare missense variant (MAF <0.010, AlphaMissense score ≥ 0.792) demonstrated or expected to be on the other allele (Table 1 and Supplementary Table 1). A total of 22 distinct missense variants were identified. Two unique missense variants occurred repeatedly in 8 unrelated families, each inherited *in trans* with different *TTNt*v, while the other 20 missense variants were unique to each family. Additionally, a further family carrying the p.(Arg25480Pro) missense variant *in trans* with a distinct *TTN*tv was previously reported by Rees and colleagues.^22^ Patients F14-P1 and F25-P1 each carry two candidate missense variants *in cis*, with both showing high AlphaMissense scores. Segregation was performed in all cases except one (F4), and the truncating and missense variants were confirmed to be inherited *in trans*. Although parents of patient F4 were unavailable for segregation analysis, the two variants are predicted to be *in trans* based on gnomAD data (Variant Co-Occurrence/Phasing tool). All 22 missense variants are found in regions of titin predicted to be folded, which make up ∼78% of the total protein sequence. Analysis of the location of the WT amino acids missense-mutated in patients showed those mutated to non-proline residues were buried in the core of their domain, while residues mutated to proline were found on both the surface and the core of domains (Figure S5, Additional File 2).

A total of 26 unique likely pathogenic/pathogenic *TTN*tv – 23 frameshift and 3 splicing variants – were identified. Both missense and truncating variants are distributed across the majority of the *TTN* gene, from exon 2 (the first coding exon) to exon 361.

Twenty-one patients (belonging to families F1-F6, F9, F10-F21) showed a congenital onset, with the first signs identified at birth or in early infancy. In these patients, the age at last evaluation varied from under 1 year to 17 years. Those patients all displayed generalized hypotonia, motor delay, and other variable features, such as pectus excavatum, scoliosis, myopathic facies, minor facial dysmorphisms, dysphagia, and respiratory insufficiency. Patient F9-P1 presented the most pronounced, syndromic-like craniofacial dysmorphisms, including a prominent nose with a broad nasal base and deviated septum, short philtrum, mandibular prognathism, maxillary hypoplasia, high-arched palate, and widely spaced teeth. He carries the p.(Ser8347Pro) variant, with a MAF of 4.027 × 10□□.

Of the 19 patients diagnosed with a congenital myopathy, five (26%) showed significant improvement after birth, as indicated by an asterisk in Table 2. All individuals, except F10-P1and F17-P1— who were in the 0–5 and 6–10 year age ranges at last follow-up —are able to walk independently without assistive devices. F10-P1 walks with support, consistent with a moderate phenotype, but can also ambulate independently for short distances. None of the individuals require ventilatory or other systemic support. All five patients carry one of the two *TTN* variants located in an exon with a PSI decreasing in skeletal muscles from prenatal to postnatal stage. The two oldest individuals in the cohort, F19-P1 and their younger sibling F19-P2, both teenagers, exhibit a stable disease course and are able to ambulate with very minimal functional limitation. They carry a splicing variant in exon 184, predicted to result in donor-site loss. This exon has low expression levels in skeletal muscle both prenatally and postnatally, with a PSI of 21% –22% respectively. The variant was found in trans with a missense variant located in a constitutively expressed exon.

Four patients (F22-F25) presented with childhood-onset symptoms, from early childhood to early adolescence. F22-P1 presented with a right-sided high-stepping gait in late childhood, which worsened over the following years with distal weakness also on the left. By mid-adolescence, he showed right-sided weakness affecting the tibialis anterior, extensor hallucis longus, neck flexors, and finger extensors, along with mild contractures of the right tibialis anterior and elbow.

F23-P1 experienced muscle stiffness and fatigue during exercise since late childhood. The MRI revealed early degenerative changes, predominantly in the medial gastrocnemius, and signs of early oedema in other calf muscles. Creatine kinase (CK) levels were markedly elevated (8059 U/L). Muscle biopsy, performed during his teenage years, from gastrocnemius, revealed fibers of variable shape and markedly variable size, including numerous hypertrophic and split fibers, atrophic fibers, and clusters of regenerating fibers. No inflammatory cell infiltration was observed. Occasional necrotic fibers were also observed, and many fibers show minicores.

F24-P1, who was a young man at the last evaluation, reported distal lower limb progressive weakness, with difficulty lifting his feet when climbing stairs. On examination as a child, he was found to have foot dorsiflexion weakness first, followed then by proximal weakness, scapular winging and distal phalanges contractures of both hands. He also showed mild facial involvement, with asymmetry of facial muscles and inability to puff one cheek. CK levels were slightly elevated (410 IU/L). He had mild restrictive respiratory impairment and recently noted arrhythmia.

Patient F25-P1 presented with disease onset from early childhood. According to the parents, she did not have neonatal hypotonia. Independent walking was achieved within the normal developmental range. The patient then developed proximal lower limb weakness, with slow progression and some difficulties climbing stairs. CK levels were normal. Muscle MRI showed muscle MRI showed a pattern highly suggestive of titin-related limb-girdle muscular dystrophy type R10 (LGMD R10), with dystrophic changes in the obturator and selective involvement of the rectus femoris, vastus intermedius, semitendinosus, and, to a lesser extent, vastus medialis and biceps femoris, while the adductor magnus was relatively spared. In the lower leg, the tibialis anterior and soleus were more affected than the gastrocnemius medialis and peroneus.

Three patients sharing the same missense variant, F7-P1, F8-P1, F8-P2, had an adult onset, although with different pattern of muscle involvement between the two families.

At the most extreme end of the late-onset spectrum, F26-P1 presented with an axial myopathy and bent spine in her seventies; on neurological evaluation, mild proximal limb girdle involvement was also detected.

### Recurring variants

Seven patients from six different families from various regions across Eastern and Southern Europe (F1-F6) carry the missense variant p.Gln7023Pro in compound heterozygosity with a *TTN*tv, which is different in each family (Table 1). p.Gln7023Pro has an AlphaMissense score of 0.81 and a MAF of 0.000005578 with only 9 alleles observed in the European population, and no reported homozygous carriers in the gnomAD database. All the patients displayed a very similar congenital myopathy phenotype, with a similar disease course. General hypotonia and muscle atrophy from birth, scoliosis, respiratory insufficiency, myopathic facies and a low weight for age were reported. All parents were reported to be healthy.

Muscle biopsies from F1-P1 and F1-P2 (both from vastus lateralis) showed common features, including variation in fibre size, increase in endomysial connective tissue, internally placed nuclei, and granulous basophilic material inside, as displayed in Figure 3 and in Figure S1, Additional File 2.

**Figure 3.**
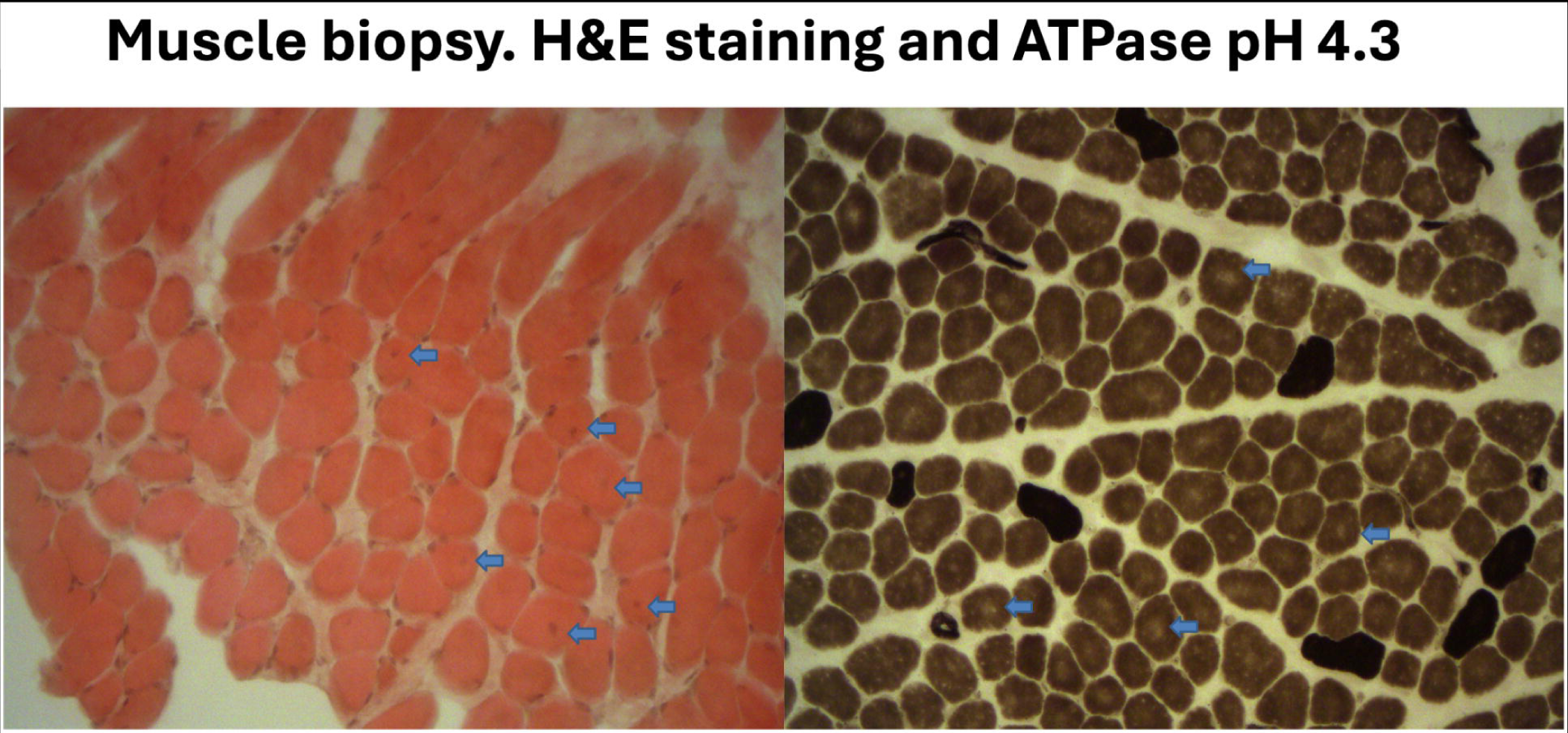
Muscle biopsy from the quadriceps (vastus lateralis). Hematoxylin and eosin staining (left) shows fiber size variability, internally displaced nuclei (arrows), and mild endomysial fibrosis. ATPase staining at pH 9.4 (right) reveals type 1 fiber predominance and perinuclear clearing (arrows).

We were able to retrieve a list of all *TTN* variants, including polymorphisms, identified in F1, F2, F5 and F6 carrying variant p.Gln7023Pro. Forty-five shared variants across the gene were identified in families F1, F2, F5 and F6 (Additional File 3). Regarding F4-P1, the phase was not available, and we were able to collect only pre-filtered data, including four missense variants, one of which was p.Gln7023Pro. Notably, F4-P1 shared two of the 45 variants found in the common haplotype with families F1, F2, F5 and F6. These two variants have gnomAD MAFs of 0.018 and 0.014, respectively. Overall, these data suggested the presence of a common haplotype among the five families, consistent with inheritance from a shared European ancestor.

Variant p.(Gln4829Pro) has a MAF of 0.0001909, with 308 alleles found mainly in the European population, and no reported homozygous carriers in the gnomAD database. It was found in three patients with adult onset from two different European families, F7 and F8, carrying *in trans* TTNtv respectively in the I-band and A-band. Patient F7-P1 showed symmetrical scapular winging, facial weakness, and distal lower and upper limb involvement with hand muscle atrophy. On muscle biopsy, central loss of NADH staining in muscle fibers was observed, sometimes with core-like lesions, with granulous basophilic material inside (Figure S2, Additional File 2). Two siblings, F8-P1 and F8-P2, showed facial weakness and predominantly proximal involvement of the lower limbs. MRI of the lower limbs shows selective fatty replacement predominantly affecting the posterior thigh and calf muscles, with relative sparing of anterior compartments (Figure S3, Additional File 2).

### Disease-linked missense variants prevent soluble expression of titin domains in bacteria

Three of the rare missense variants, collectively found in 11 of the 30 patients presented here, were studied in vitro to assess the effect of the variants on domain stability and aggregation. Titin Ig-31, Ig-54 and Ig-68, and their variants Q4829P, Q7023P and S8347P, respectively (Fig. S4, Additional File 2), were expressed in bacteria and their solubility was assessed by lysing the cells, separating the soluble and insoluble material and determining in which fraction they were found. Western blotting using an antibody that recognised the recombinant proteins’ His-tag showed that while all domains were expressed and the three WT domains were found in the soluble fraction, all three variant domains were absent, demonstrating that the variants prevented the soluble expression of their domain (Figure 4).

**Figure 4.**
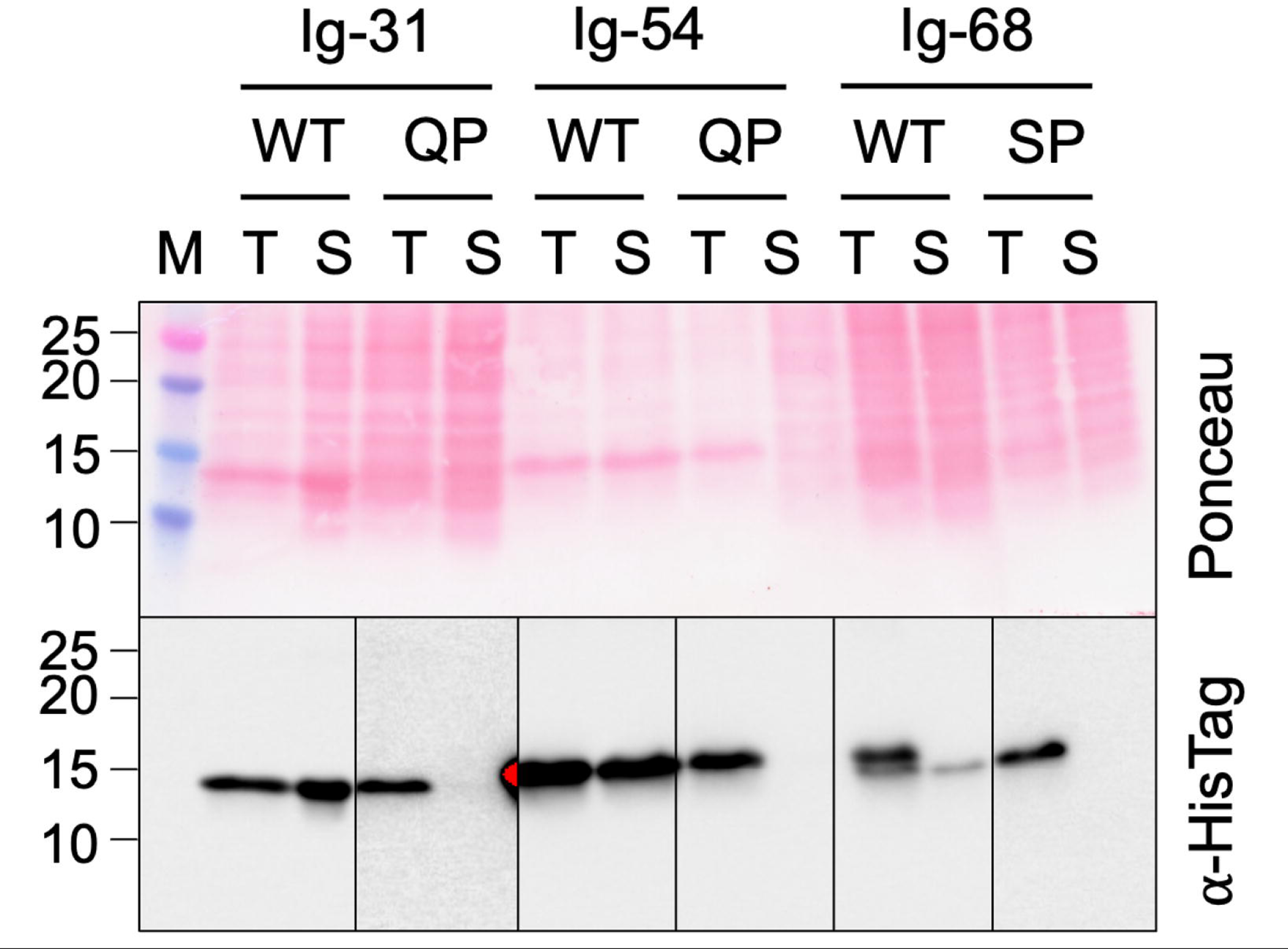
Assessment of solubility of WT and missense variant-containing titin domains. Western blot probing presence of His-tagged titin domains Ig-31 WT and Q4829P, Ig-54 WT and Q7023P and Ig-68 and Q7023P in the soluble fraction of bacterial expression lysate. T = total lysate, S = soluble fraction, M = molecular weight marker.

### Disease-linked missense variants alter the expression pattern of titin domains in COS-7 cells

GFP-tagged constructs of the same WT and variant titin domains were expressed in COS-7 cells, and the pattern of expression and co-localisation with a marker of proteostasis were assessed using fluorescent microscopy. Confocal micrographs of the transfected cells showed that the WT titin domains had a diffuse expression pattern throughout the cell, with stronger localisation to the nucleus. In contrast, the variant domains had a largely punctate, presumably aggregated appearance, and were excluded from the nucleus (Figure 5A, Figure 6). We quantified these differences by analysing widefield images of transfected cells and measuring the Jaccard Index of nuclear overlap with the whole cell mask; all three WT-variant pairs showed a significant difference in their cell expression pattern (Figure 5B, Fig. S6, Additional File 2).

**Figure 5.**
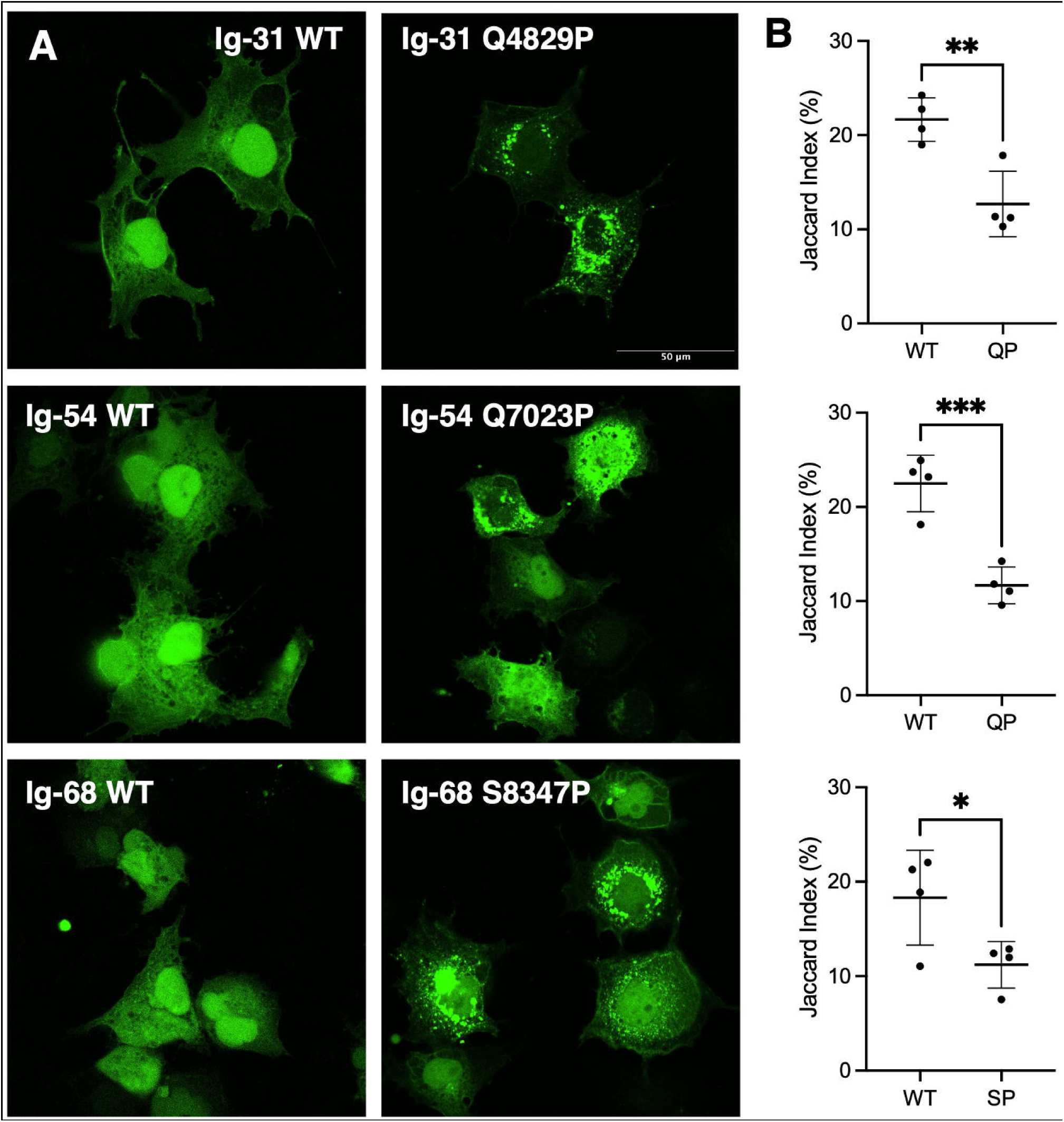
Expression of WT and missense variant-containing titin domains in COS-7 cells. **(A)** Confocal microscopy images showing expression of GFP-tagged titin domains Ig-31 WT and Q4829P, Ig-54 WT and Q7023P and Ig-68 WT and S8347P. **(B)** Quantification of widefield fluorescence microscopy images of the expression pattern of GFP-tagged titin domains. A lower Jaccard index correlates with a more punctate expression pattern. * = *P* < 0.05, ** = *P* < 0.005.

**Figure 6.**
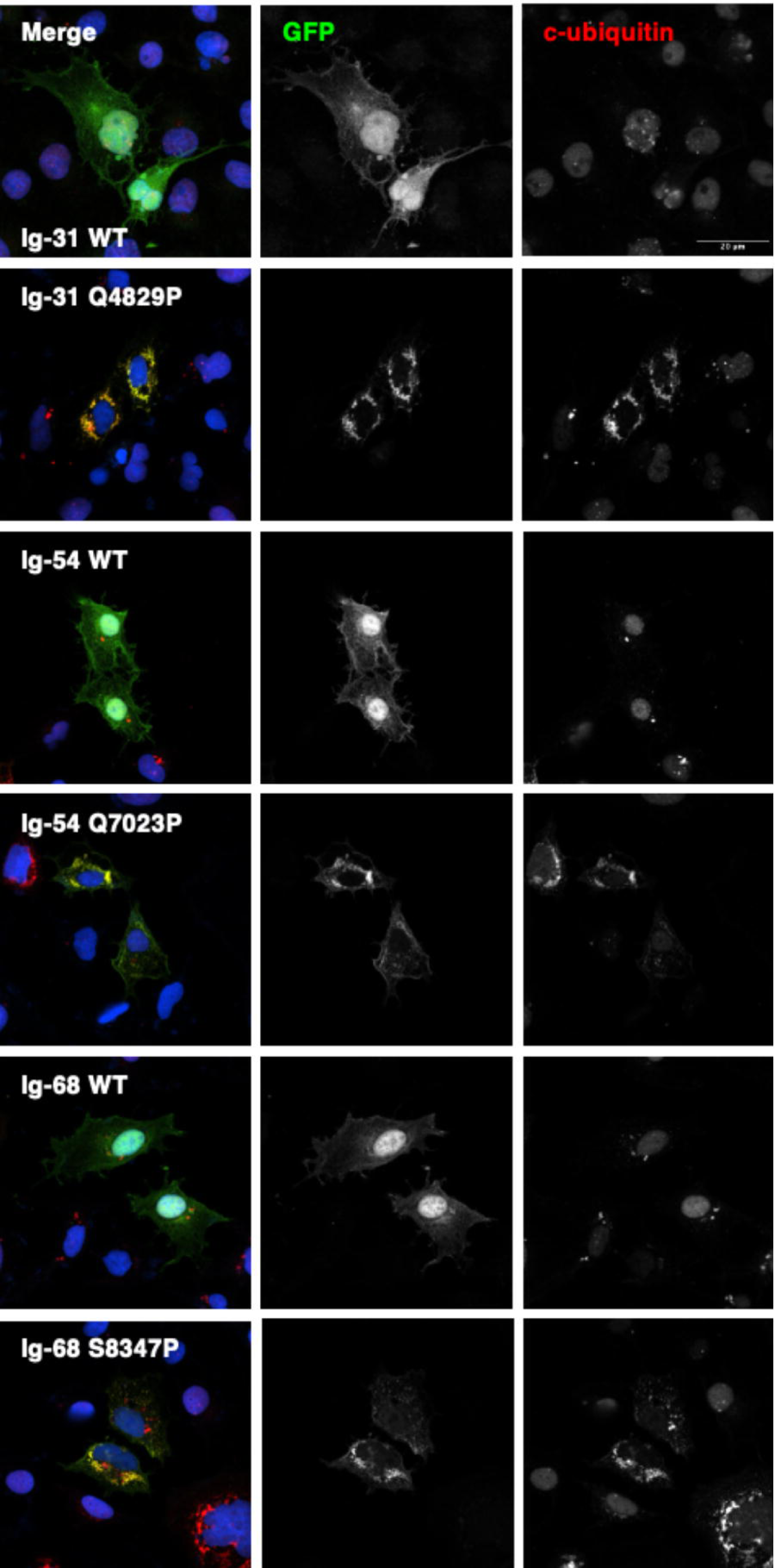
Localisation of conjugated ubiquitin in COS-7 cells expressing titin domains. Confocal microscopy images showing expression of GFP-tagged titin domains Ig-31 WT and Q4829P, Ig-54 WT and Q7023P and Ig-68 and Q8437P (GFP, green) co-stained with an antibody that recognises conjugated ubiquitin (c-ubiquitin). Regions of co-localization of GFP-tagged protein and conjugated ubiquitin are shown in yellow. Nuclei are shown in blue.

As we presumed the punctate expression was due to the missense variant domains aggregating as a result of the variant preventing correct protein folding, we co-stained the fixed cells with an antibody that recognises conjugated ubiquitin (c-ubiquitin), a protein that is enzymatically linked to misfolded proteins targeted for degradation via the ubiquitin-proteosome system or autophagy pathway (Figure 6). While there was little co-localisation of GFP and c-ubiquitin for the WT domains, c-ubiquitin frequently co-localised with the variant-containing domains and displayed the same punctate appearance, suggesting that the variant domains are misfolded and are being targeted for degradation.

## Discussion

Our work collected evidence that rare missense variants can contribute to recessively inherited titinopathies, presenting with a broad phenotypic spectrum. To support a more systematic and scalable approach to variant interpretation, we also aimed to develop a practical framework for assessing *TTN* missense variants in a diagnostic setting, integrating in silico prediction tools, functional assays and clinical data.

### AlphaMissense and MAF

In our cohort, AlphaMissense proved to be a valuable tool to support the application of the ACMG criterion PP3 for the missense variants identified, providing evidence of pathogenicity based on in silico predictions. Although, as a standalone criterion, PP3 is insufficient to classify a variant as likely pathogenic or pathogenic, AlphaMissense scores can play a crucial role in guiding the interpretation of the vast number of missense variants in *TTN*. This trend is further evidenced by the rapid escalation in the application of AlphaMissense to the interpretation of missense variants in clinically relevant genes, such as *BRCA1* and *TP53*.^32,33^ In the current study, we investigated how these considerations specifically translate to the *TTN* gene and to our cohort of unsolved patients. The overall scores for each amino acid change in TTN have been summarized in Figure 2B, which aims at facilitating the decision of whether to prioritise a rare missense variant in the absence of other evidence. MAF and the biochemical properties of any given amino acid are among the primary features considered by AlphaMissense.^24^ For instance, tryptophan’s large, hydrophobic structure and its key position within Ig and FN3 domains—where it helps stabilize the hydrophobic core—make its substitution particularly disruptive to protein structure.^34,35^ In our cohort, indeed, there are 3 variants from tryptophan and one to tryptophan. Similarly, proline, due to its rigid cyclic structure, imposes conformational constraints that can severely impact protein folding and function. Notably, recent studies have emphasized the importance of proline and other residues in maintaining the structural integrity of Ig-domains.^36^ Besides proline, also arginine, despite its markedly different structure, can disrupt local protein folding due to its basic and bulky side chain, especially when replacing an amino acid that contributes to the core of titin’s Ig and Fn3 domains. In our cohort, nine changes are to proline and seven to arginine, making them the two most frequent resulting amino acids.

However, regardless of the MAF, our analysis showed that approximately 40% of all possible missense variants in titin have an AlphaMissense score of between 0.170 and 0.791 (“indeterminate”) which doesn’t allow us to apply either criterion PP3 or BP4 to these (Figure 2A). All missense variants in our cohort have a MAF < 0.0001, with the exception of three cases. The most frequent variant is p.Leu24049Pro, with a gnomAD MAF of 0.0012, 4 homozygous individuals in gnomAD, and an AlphaMissense score of 0.989. Like most others, this variant remains classified as a VUS, and in the absence of functional studies we lack additional evidence to support its pathogenicity. However, it is worth noting that the phenotype in F27-P1 is very late-onset and falls within the mild end of the spectrum; axial myopathies in elderly patients are often underdiagnosed, and no precise prevalence estimates currently exist.

The second most frequent missense variant is p.(Gln4829Pro), which has a MAF of 1.91 × 10CC. We have shown in vitro that this variant affects domain folding and alters the expression pattern. The variant is found in families F7 and F8, which both show an adult-onset phenotype. The third, p.(Val27761Leu) was found in patient F16-P1, who had a congenital onset but carries the variant *in cis* with another missense variant, which is rarer, but has a similarly high AlphaMissense score. Importantly, no congenital case in our cohort involves a missense variant with MAF > 0.0001, which is in line with expectations considering the population prevalence of congenital myopathies.

Among the 3,086 missense variants predicted as possibly pathogenic by AlphaMissense and found in gnomAD, 96 were observed in homozygosity in at least one individual, with MAFs ranging from 0.000004357 to 0.0284. Twenty-one had a MAF > 0.001 and 4 are results of a polymorphism (MAF >0.01), possibly reflecting AlphaMissense overestimation of a variant’s pathogenicity due to domain redundancy and structural similarity in the TTN protein. Filtering out rare variants with a few homozygotes is less straightforward, however, as some may be hypomorphic and clinically relevant only *in trans* with a *TTN*tv, particularly in high-PSI exons.

Nonetheless, after removing variants observed in homozygosity, we estimated a cumulative frequency of high-scoring missense alleles that— combined with a 0.05% prevalence of *TTN*tv in constitutively expressed exons (i.e., high-PSI *TTN*tv only), and assuming 50% chance of trans configuration—would result in ∼1 in 9,150 individuals carrying both a *TTN*tv and a missense variant of possible clinical significance. While this high prevalence may partly reflect underdiagnosed or mild late-onset cases, our data suggest that AlphaMissense tends to overestimate pathogenicity in *TTN*, and that variant interpretation in this gene requires additional considerations. Overall, considering these findings in the context of our current knowledge, for severe, early-onset myopathies, a MAF cut-off of < 0.0001 appears appropriate. In milder cases with adult or late onset, filtering by MAF becomes even more challenging. As a general rule, we suggest that the higher the MAF, the stronger the other lines of pathogenicity evidence must be for a variant to be considered potentially disease-causing.

### ACMG criteria

Among the reported variants, only p.(Gln7023Pro) – functionally assessed here and identified in six unrelated families – and p.(Arg25480Pro) – previously functionally assessed and now reported in a second, unrelated family – meet the criteria for classification as likely pathogenic. This classification is based on the application of the following ACMG/AMP criteria:

– PM3 (Moderate for p.(Gln7023Pro) and Supporting for p.(Arg25480Pro)): A total of 1.25 points were assigned for the first and 0.50 for the latter, in line with ClinGen guidelines for large genes, which recommend awarding 0.25 points per proband, considering the large size of the gene.
– PS3 (Moderate for both): Functional studies were conducted using assays which have been previously used for over ten pathogenic and benign variants, but without a formal statistical analysis of the ability to discriminate between pathogenic and benign variants.^18^
– PP1 (Strong for variant p.(Gln7023Pro) and Supporting for p.(Arg25480Pro)): Co-segregation with disease was observed across seven informative meioses in five families in the case of p.(Gln7023Pro), and across two informative meioses in two families in the case of p.(Arg25480Pro). In F1, the variant was present in two affected siblings with confirmed parental segregation. In F2, a healthy sibling was tested and carried only one of the two variants, consistent with the expected recessive inheritance.
– PP3 (Supporting for variant p.(Gln7023Pro) and Strong for p.(Arg25480Pro)): The AlphaMissense score was 0.83 and 0.99, respectively, exceeding the threshold recommended by ClinGen.
– PM2 (Supporting for both variants): “gnomAD genomes homozygous allele count = 0 is less than 2 for AD/AR gene *TTN*, good gnomAD genomes coverage = 31.8” and “Variant not found in gnomAD genomes”.
– PP4 (Supporting strength for both variants): The clinical phenotype and disease progression are consistent with recessive congenital titinopathy in all cases. Moreover, muscle biopsy findings are in line with those reported in other confirmed congenital titinopathy cases.
– BP1 (Supporting strength) was manually deactivated for both variants. Although historically *TTN* has been associated exclusively with truncating variants, this is largely due to a bias in variant interpretation and reporting, with limited systematic study of missense changes. Importantly, missense variants located in specific exons of *TTN* have already been reported as pathogenic (exons 344, 364). Therefore, the assumption underlying BP1—that missense variants in this gene are unlikely to be disease-causing—is not valid.

In the case of p.(Gln7023Pro) and p.(Arg25480Pro), the experimental assays provided variant-specific functional evidence, with direct clinical implications—namely, a confirmed genetic diagnosis for affected individuals and the possibility of prenatal or preimplantation testing for at-risk families. Additionally, RNA-seq performed on individual F1-P2 did not reveal any splicing alterations, further reinforcing the interpretation of this specific missense variant. Without such assays, it would have been more appropriate to consider the shared allele observed in the six families as pathogenic, rather than attributing pathogenicity directly to the variant itself, since the possibility of another undetected variant in cis could not be excluded. This issue is particularly relevant for other missense variants identified in our cohort, for which no functional assays or second-tier analyses (such as RNA-seq or long-read sequencing) are available. In F25-P1, muscle MRI revealed a pattern characteristic of titinopathies (LGMD R10). RNA sequencing did not reveal any cryptic abnormalities. Based on these findings, the following ACMG/AMP criteria were applied: PP4 (Supporting), PM2 (Moderate; variant absent from gnomAD), and PP3 (Strong; Alphamissense score of 0.9997 with full concordance across in silico prediction tools). Although this combination would classify the variant as likely pathogenic, we found it more appropriate to suggest the pathogenicity of the allele, as the variant has been identified in a single patient and in cis with a second rare missense change, also predicted to be deleterious (Alphamissense score: 0.9851), as reported in Table S2, and we cannot accurately determine the specific contribution of each variant. In F17-P1, carrying in-trans variants p.(Trp9934Arg) and p.(Arg30557Ter), long-read sequencing (Nanopore) was performed and ruled out the presence of other variants of interest, including structural variants and copy number variations (CNVs). In the remaining cases, the possibility of undetected in-cis variants—such as additional VUS, non-canonical splicing events, or CNVs—cannot be confidently excluded and should be acknowledged as a limitation.

Notably, in large and complex genes such as *TTN*, segregation across multiple informative meioses—while valuable—typically provides strong evidence for the pathogenicity of the allele as a whole, rather than of the specific variant. To establish variant-level pathogenicity, additional supporting evidence is required. This may include the identification of unrelated cases carrying the same variant on distinct haplotypes, which is unlikely for extremely rare variants, or functional assays specifically addressing the variant’s effect. Additional strategies, such as RNA-seq, long-read sequencing, or other second-tier genomic approaches, can help exclude the presence of cryptic *cis*-variants and further strengthen the interpretation of rare missense changes.

### Functional assays

Our functional studies demonstrate that p.(Gln4829Pro), p.(Gln7023Pro) and p.(Ser8347Pro) impair protein folding and stability, and promote protein aggregation in vitro (Figure 5, Fig. S7, Additional File 2). These findings are consistent with the known role of proline as a β-sheet breaker, due to it disrupting local folding on account of its rigid structure and its inability to act as a main chain hydrogen bond donor. To gain additional evidence, the surface accessibility of the WT amino acid mutated in patients was determined, and defined as either a surface-exposed or core residue (Fig. S6, Additional File 2).^26^ All not-to-proline mutations occur at buried sites, suggesting a disruption of domain stability. In contrast, some substitutions to proline affect more exposed residues but still appear pathogenic, likely due to proline’s structural rigidity and its tendency to disrupt local secondary structure, even when positioned on the domain surface (Fig. S6, Additional File 2:).^29,30^ All missense variants in the cohort are located within structured domains of TTN (i.e., Ig, Fn3, or kinase domains). This may reflect the greater pathogenic potential of variants in these conserved regions, but it is also important to acknowledge a potential bias: AlphaFold is less accurate in predicting unstructured regions, and therefore AlphaMissense may not provide reliable predictions for variants in those regions.

In addition, while functional assays do provide important mechanistic insights, several limitations on their use must be considered. Titin is an exceptionally large and modular protein, and such assays are performed on a single domain. As a result, they may not accurately recapitulate the structural and functional complexity of the full-length protein *in vivo*, including domain–domain interactions, ligand interactions, and tissue-specific regulation. Moreover, these studies are time– and resource-intensive, making them impractical for high-throughput applications across the vast number of *TTN* missense variants observed in patients. In the future, scalable and standardized functional assays will be essential to enable broader variant characterization in critical *TTN* regions. At the same time, it will be important to carefully assess the weight of these assays in variant classification frameworks.

Lastly, no pathogenic variants in other muscle disease genes suggestive of digenic inheritance were detected; although such mechanisms appear unlikely—particularly in cases with recurring and functionally assessed variants—we cannot entirely rule out unknown forms of digenic inheritance.

## Conclusions

Establishing the pathogenicity of missense variants in *TTN* remains challenging, even when functional assays are available, as is accurately predicting their *in vivo* effects. In this study, we demonstrated that additional insights can nevertheless be gained through the in-depth analysis and collection of clinical cases, as exemplified by the emblematic case of a novel haplotype defined by the now likely pathogenic p.(Gln7023Pro) variant. Also, we suggested that aggregation and disruption of the β-sheet secondary structure in Ig-domains may represent a potential mechanism of pathogenicity for selected missense changes. However, genotype–phenotype correlations cannot be attempted yet: the clinical spectrum is broad, as are the titin domains involved, and many other cases would be required. Novel tools, such as AlphaMissense, can aid in the interpretation of *TTN* missense variants, although their tendency to overestimate pathogenicity and other limitations should be considered. We recommend that missense variants with potential pathogenicity based on in silico predictions, low MAF, or other supporting evidence be systematically reported in clinical records and internal databases when found *in trans* with a *TTN* truncating variant in patients with a compatible myopathic phenotype. Whenever feasible, these findings should also be shared across international variant repositories and neuromuscular networks to promote data integration and support future reclassification efforts.

## Data Availability

All data produced in the present work are contained in the manuscript

http://psivis.it.helsinki.fi:3838/TTN_PSIVIS/

## Abbreviations

TTN: Titin
Fn3: Fibronectin type III-like
Ig: Immunoglobulin-like
MAF: Minor Allele Frequency
HMERF: Hereditary Myopathy with Early Respiratory Failure
TMD: Tibial Muscular Dystrophy
TTNtv: TTN truncating variant
PSI: Percent Spliced In
DCM: Dilated Cardiomyopathy
ACMG: American College of Medical Genetics and Genomics
gnomAD: Genome Aggregation Database
CNV: Copy Number Variation
RNA-seq: RNA sequencing
WT: Wild Type
pET: Plasmid Expression Vector
His6-tag: Hexahistidine tag
pCMV_GFPC2: GFP-tagged expression vector
LB: Luria-Bertani (broth for bacterial culture)
ECL: Enhanced Chemiluminescence
HRP: Horseradish Peroxidase
GFP: Green Fluorescent Protein
COS-7: Monkey kidney fibroblast-derived cell line
PBS: Phosphate Buffered Saline
PFA: Paraformaldehyde
DAPI: 4′,6-diamidino-2-phenylindole
VUS: Variant of Uncertain Significance
LGMD R10: Limb-Girdle Muscular Dystrophy Recessive 10

## Ethics declaration

This study involves human participants and ethical approval for this study falls under HUS/16896/2022 by the HUS Ethics Committee. The study of the samples was performed in accordance with the Declaration of Helsinki. Participants gave informed consent to participate in the study before taking part.

## Acknowledgements

We thank all patients and families participating in this study.

## Supplementary Information

**Additional File 1.** List of rare *TTN* missense variants considered in this study. AlphaMissense scores and MAF from gnomAD are reported.

**Additional File 2.** Supplementary clinical, histological, and functional data.

**Additional File 3.** Shared *TTN* haplotype in families carrying the p.(Gln7023Pro) variant. List of *TTN* variants defining a common haplotype identified in multiple unrelated families carrying the likely pathogenic variant p.(Gln7023Pro).

