## Additional File 2 for "A comprehensive framework for the interpretation of *TTN* missense variants"

**Additional File 2. Supplementary clinical, histological, and structural variant data**

**Index**

Figure S1. Muscle biopsy from Patient F1-P1.

Figure S2. Muscle biopsy of Patient F7-P1.

Figure S3. Muscle MRI of Patient F8-P1.

Figure S4. Experimental or computationally predicted structures of titin domains (Ig-31, Ig-54, and Ig-68) studied in vitro

Figure S5. Quotient surface accessible surface area q(SASA) of wild type (WT) amino acids mutated to either proline or any other amino acid in patient cohort

Figure S6. Widefield fluorescence microscopy images of COS-7 cells expressing GFP-tagged WT and missense variant-containing titin domains.

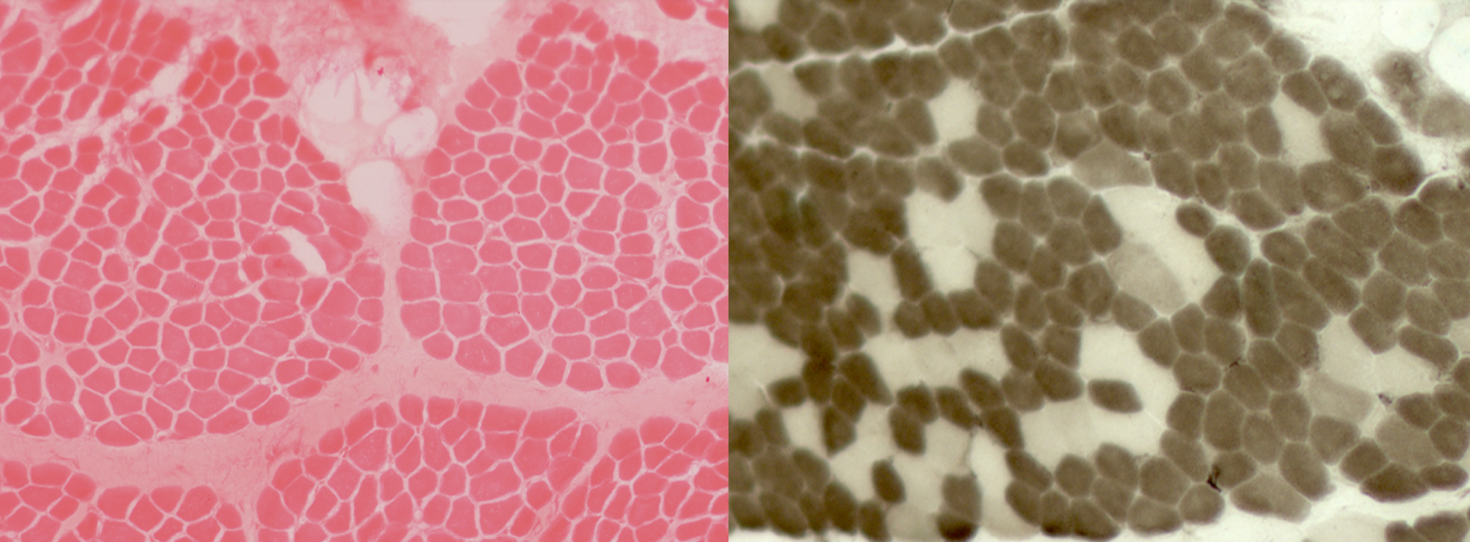

**Figure S1.** Patient F1-P1, muscle biopsy. Vastus lateralis, performed in infancy.

Variability in muscle fibre calibre due to the presence of some atrophic fibres and a slight increase in endomysial connective tissue. ATPase pH 4.3 shows predominance of type 1 fibres.

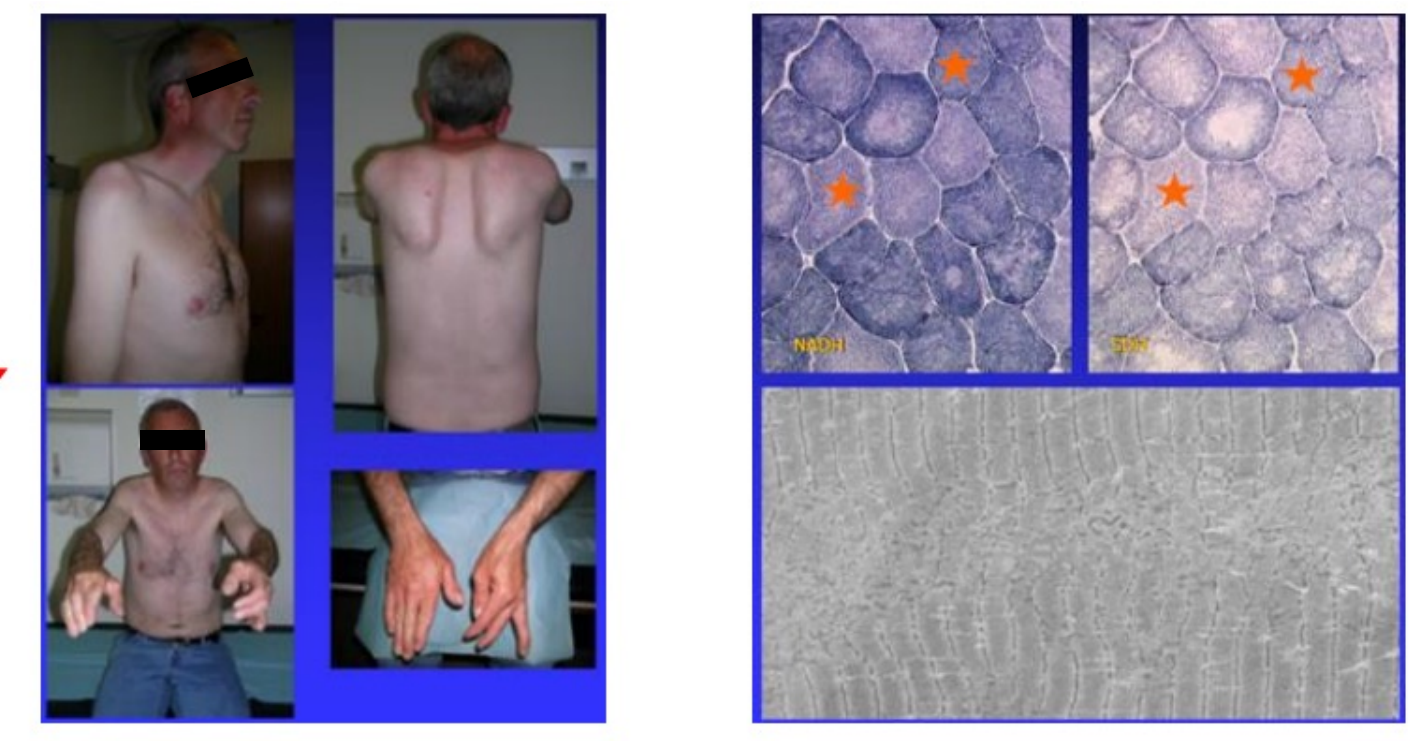

**Figure S2.** Patient F7-P1, showing distal involvement, scapular winging. Muscle biopsy (NADH staining) showing scattered atrophic fibers with structural disorganization and minicores (indicated by stars).

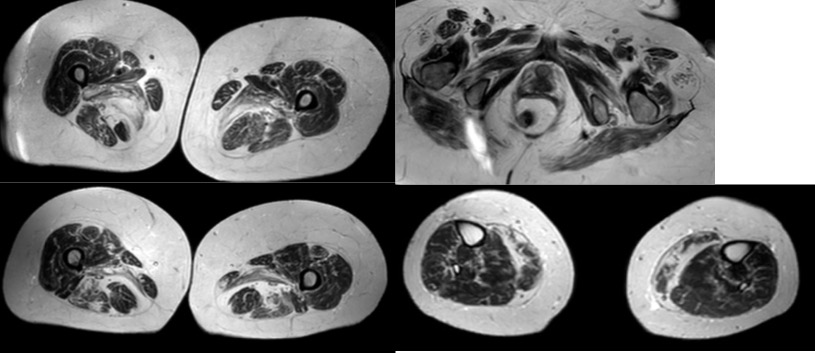

**Figure S3.** Patient F8-P1, muscle MRI. Marked fatty replacement and atrophy of posterior thigh and calf muscles, with relative sparing of anterior compartments.

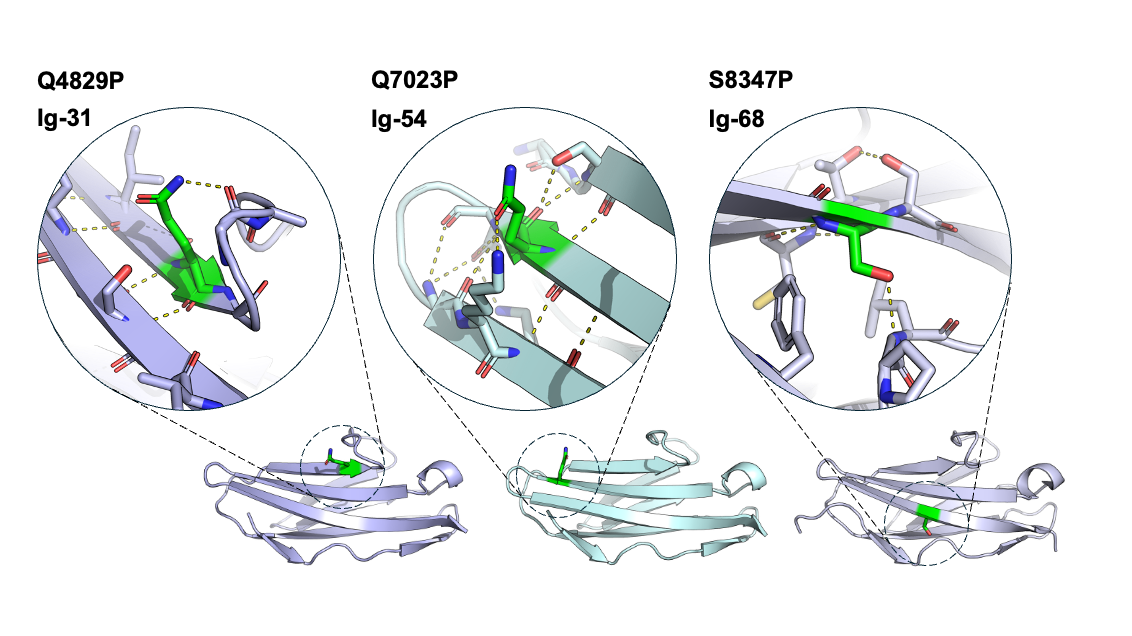

**Figure S4**. Experimental or computationally-predicted structures of titin domains studied in vitro showing the atomic environment of residues mutated in patient missense variants to proline. The amino acids mutated in patients are shown as green sticks, with surrounding main-chain and side-chain atoms of interest also shown as sticks. Polar contacts between these residues are shown as dashed yellow lines. Ig-31 and Ig-54 are AlphaFold-predicted models downloaded from TITINdb. Ig-68 is from an experimental structure, with PDB code 3B43. Each of the three residues mutated in patients are at a different structural position, but all are located on a beta strand. Proline is typically incompatible with beta strands – it is a ”beta-breaker”.

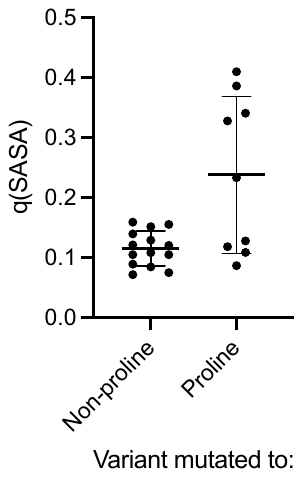

**Figure S5.** Quotient surface accessible surface area q(SASA) of amino acids mutated to either proline or any other amino acid in patient cohort, calculated from predicted or experimental domain structures. Amino acids with q(SASA) values above or below 0.3 are defined as surface or core residues, respectively. q(SASA) value retrieved from TITINdb (<https://titindb.kcl.ac.uk/TITINdb/> ).

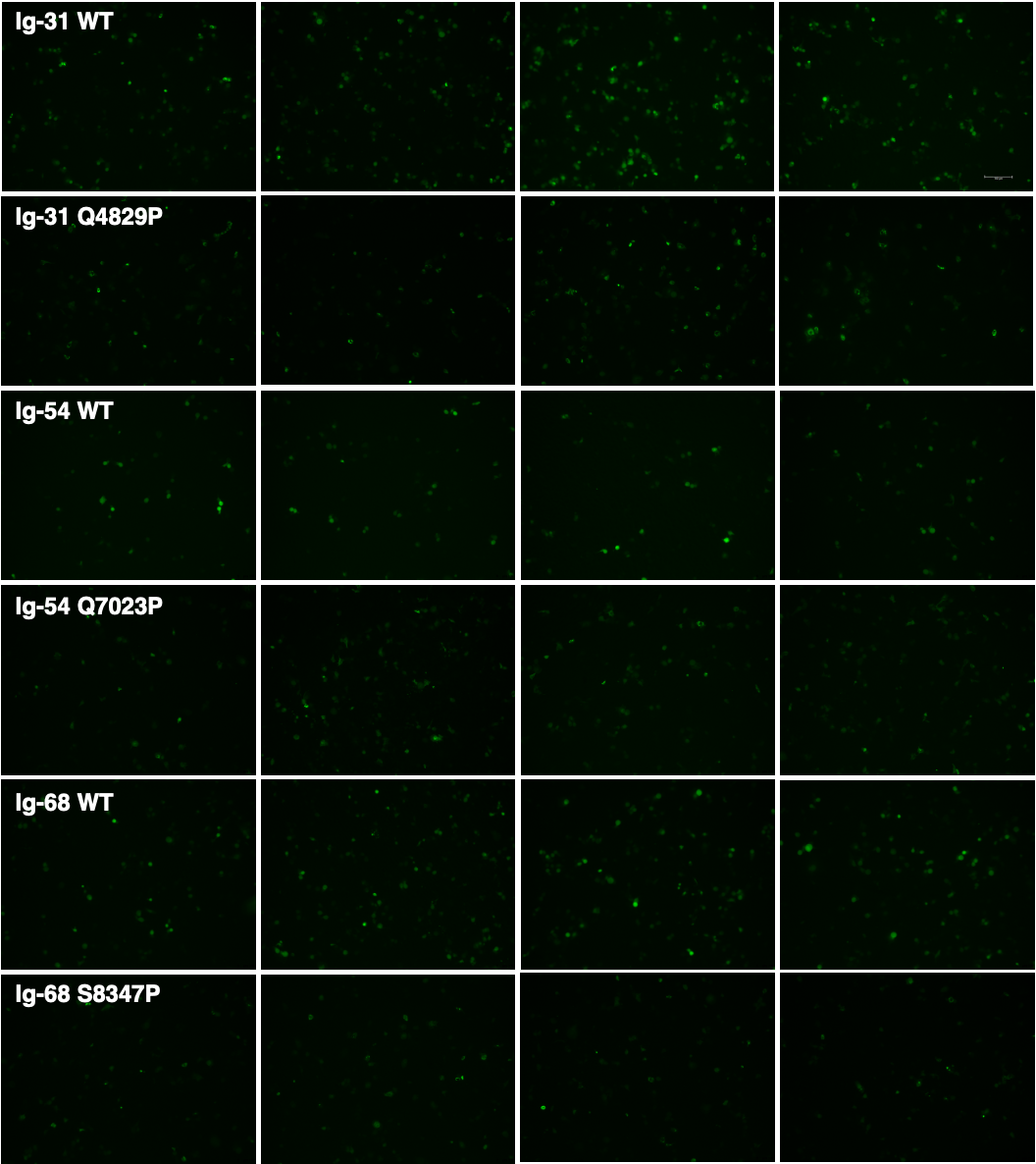

**Figure S6.** Widefield fluorescence microscopy images of COS-7 cells expressing GFP-tagged WT and missense variant-containing titin domains. These images were used for the images analysis shown in figure 5B. Scale bar = 150um.
